# Stratifying Eating Disorders with Clustering: From Diagnosis to Phenotypic Diversity

**DOI:** 10.1101/2025.08.25.25334344

**Authors:** Andrea Zanola, Louis Fabrice Tshimanga, Valentina Meregalli, Angela Favaro, Manfredo Atzori, Enrico Collantoni

## Abstract

Psychiatric disorders, including eating disorders (EDs), are characterized by heterogeneity that limits diagnostic accuracy and treatment personalization. Precision psychiatry calls for tools to identify data-driven phenotypes beyond traditional categories. In a large cohort (N=809), we applied an unsupervised clustering pipeline to self-report and clinical variables to uncover ED subgroups. Repeated Spectral Clustering revealed four phenotypes: two aligned with DSM-5 diagnoses (anorexia nervosa restricting-type; bulimia nervosa), and two diagnostically mixed clusters, one characterized by higher psychopathology and trauma exposure, the other by prolonged illness duration. These clusters showed distinct profiles and outcomes. The higher predictability of data-driven clusters compared to DSM-5 categories suggests that unsupervised stratification may offer a complementary perspective on relevant heterogeneity. Our findings highlight how computational phenotyping can advance precision psychiatry by revealing meaningful patterns, informing prognosis, and guiding personalized interventions. This approach may help bridge the gap between clinical presentations and the need for stratified, patient-centered care.

## 1 Introduction

Psychiatric disorders are characterized by substantial clinical heterogeneity, posing challenges for diagnosis, prognosis, and treatment planning. Eating disorders (EDs), including anorexia nervosa and bulimia nervosa, exemplify this complexity, exhibiting diverse trajectories, comorbidities, and levels of treatment response [1]. Although these conditions share several elements of psychopathological and behavioral homogeneity, as outlined in the Diagnostic and Statistical Manual of Mental Disorders, Fifth Edition (DSM-5) [2], patients with EDs often demonstrate a wide range of symptom profiles, comorbidities, and clinical trajectories that extend beyond conventional diagnostic subtypes [3]. This inherent variability complicates efforts to develop targeted therapeutic interventions, accurately predict outcomes, and refine clinical strategies.

In this context, increasing attention has been devoted to identifying more nuanced and empirically defined subgroups—or phenotypes—within clinical populations with EDs [4, 5]. To this end, data-driven methods such as network modeling and cluster analysis have been employed to examine a broad range of psychological variables, personality dimensions, demographic factors, and clinical indicators [6]. These approaches aim to reveal distinct patterns that transcend conventional nosological frameworks, placing emphasis on how dimensions beyond core EDs symptoms can critically shape the clinical picture. Recent evidence suggests that, within EDs samples, discernible groups emerge on the basis of factors such as disordered eating behaviors, body image disturbances, interpersonal functioning, and underlying temperament and personality traits [7]. In addition, a growing body of research emphasizes the importance of examining symptoms beyond the confines of rigid diagnostic categories, underscoring the critical role of transdiagnostic mechanisms in EDs. These mechanisms may influence not only the underlying pathophysiology but also the persistence, clinical complexity, and treatment responsiveness of these conditions. This evidence appears to align with the well-established theoretical view that conceptualizes EDs within a transdiagnostic framework, which underpins cognitive-behavioral therapies for these conditions [8].

In recent years, machine learning (ML) has emerged as a powerful and transformative approach within the psychiatric context, enabling the analysis of complex, high-dimensional data and facilitating the discovery of latent patient subgroups [9]. This capability is particularly relevant for heterogeneous disorders such as anorexia nervosa and bulimia nervosa, where conventional diagnostic categories often fail to capture the underlying variability and where the boundaries defined by diagnostic criteria do not prevent high rates of transdiagnostic migration and the emergence of atypical phenotypes [10]. Unsupervised ML techniques, such as clustering, are especially promising as they can identify subgroups without relying on predefined categories, offering a more flexible and data-driven framework for understanding EDs subtypes. Despite this promise, existing ML applications to EDs populations have been somewhat fragmented [11]. Previous research has pursued a variety of goals, from early detection of those at risk to predicting treatment responses, using a broad range of methodologies, from basic regression models to advanced neural networks [12]. For instance, Rosenfield and Linstead [12] employed cluster analysis to examine the relationships among the Eating Disorder Examination Questionnaire, Clinical Impairment Assessment, and Autism Quotient, showcasing the potential of ML to identify vulnerability to EDs. Using a different approach, Haynos and colleagues [13] demonstrated that ML techniques outperformed logistic regression in predicting the longitudinal course of EDs. Furthermore, Juarascio *et al*.[14] emphasized the potential of just-in-time adaptive interventions, which use ML or predictive algorithms to identify moments of heightened risk for disordered eating behaviors and deliver personalized, real-time strategies to improve treatment outcomes. Notably, many of these studies remain anchored in traditional diagnostic labels, and fewer have fully explored unsupervised methods that might reveal previously unrecognized patient profiles or confirm the presence and the possible clinical utility of novel phenotypes.

The utility of ML in psychiatry is not limited to EDs. Similar approaches have already shown promise in other complex mental health conditions, such as mood disorders and schizophrenia, where data-driven subgroup identification has led to the discovery of clinically meaningful phenotypes and improved stratification for interventions [15]. In this context, specific unsupervised machine learning methods, such as Gaussian mixture models and clustering algorithms, have been used to discover distinct neuroanatomical and cognitive subtypes within schizophrenia populations [16, 17, 18], revealing subgroups with differing brain structural and cognitive patterns [19]. Similarly, in the field of depression research, ML approaches have been increasingly utilized to address the substantial heterogeneity of the disorder, which often hinders precise diagnosis and treatment. For instance, latent variable models and clustering techniques have been applied to large datasets to identify empirically derived depressive subtypes, enabling a better understanding of symptom variability and its relationship to clinical outcomes [20]. This cross-diagnostic evidence supports the application of ML-based strategies in the psychiatric context, where there remains a pressing need to enhance diagnostic precision and treatment personalization. Building on the ED-specific and cross-diagnostic work outlined above, unsupervised techniques represent a valuable tool for exploring latent phenotypic structures that extend beyond the boundaries of established diagnostic classifications. By clustering readily available psychometric and clinical variables, such models can simultaneously refine diagnostic boundaries, furnish pragmatic decision tools for clinicians, and generate falsifiable predictions about prognosis and treatment response.

Building on growing evidence that ML can reveal clinically relevant patterns in psychiatric populations, we applied an unsupervised clustering pipeline to a large cohort of individuals with EDs. The primary objectives were twofold: first, to examine whether the resulting clusters correspond to traditional DSM-5 diagnostic categories or instead reflect novel, clinically meaningful phenotypes; and second, to investigate whether these data-driven subgroups are associated with differential treatment outcomes. By addressing these aims, the study investigates whether cluster-based phenotyping can enhance clinical understanding and prognostic accuracy, thereby supporting more personalized and effective interventions.

## 2 Materials and Methods

### 2.1 Participants

The cohort under analysis consists of 809 patients admitted to the Eating Disorders Unit of Padua Hospital between 2005 and 2023. Inclusion criteria were: (1) female gender and (2) a diagnosis of acute AN or BN according to DSM-5 criteria [2] at the time of the initial clinical evaluation. Among these patients, 351 (43%) were diagnosed with the restrictive subtype of AN (AN-R), 150 (19%) with the binge-purging subtype of AN (AN-BP), and 308 (38%) with bulimia nervosa (BN). The mean age was 23.1 years (standard deviation, SD = 6.8 years), with a range between 12.7 and 57.0 years.

All subjects gave their informed written consent to the use of data in an anonymous form. The Ethical Committee of Padova Hospital approved the study (protocol n*◦* 1598P).

### 2.2 Procedure and Measures

At their first admission to the center, all patients underwent a routine baseline assessment. This included a semi-structured interview covering sociodemographic and clinical variables, as well as the completion of a series of self-administered questionnaires and scales. From the interview, the following variables were extracted: diagnosis according to DSM-5 criteria [2], age, age of onset (defined as the age at first occurrence of an eating disorder), illness duration, weekly hours of physical activity, current Body Mass Index (BMI), lowest lifetime BMI, weight suppression (defined as the difference between the highest lifetime BMI and current BMI), presence of amenorrhea, and history of either sexual or physical childhood abuse. BMI was measured in kg/m2; henceforth, the unit will be omitted for brevity. The semi-structured interview also included a checklist of 27 stressful events. Patients were asked to indicate whether they had experienced any of these events in the six months preceding the onset of the eating disorder, and to rate the subjective severity of each event on a scale from 1 (slight) to 5 (catastrophic). The total severity score was used as a variable in the analyses. At the time of discharge, each patient’s clinical status was recorded and classified as full remission, partial remission, or persistent illness. Full remission was defined as the absence of eating disorder symptoms in the last month, including restored weight (BMI *≥* 18 kg/m2), absence of amenorrhea (when not due to other medical causes), no binge eating or purging behaviors, and no severe body image disturbance. Partial remission was defined as the persistence of only one of the following symptoms: amenorrhea (not explained by other medical conditions), BMI lower than 18, presence of binge eating, purging, or severe body image disturbance. No remission (persistent illness) was defined by the presence of at least two of the aforementioned symptoms at the end of the treatment [21].

The self-administered questionnaires included:

#### Eating Disorder Inventory-1 (EDI-1) [22]

The EDI-1 is a 64-item self-report measure to assess the severity of eating disorder psychopathology. Higher scores indicate greater symptom severity. All the 8 subscales were considered in this study: drive for thinness (DR-THIN), bulimia (BUL), body dissatisfaction (BODY-DISS), ineffectiveness (INEFF), perfectionism (PERF), interpersonal distrust (INT-DIS), interoceptive awareness (INT-AW), and maturity fears (MAT-FEAR).

#### Temperament and Character Inventory (TCI) [23]

The TCI is a 240-item self-report questionnaire designed to evaluate four dimensions of temperament—Novelty Seeking (NS), Harm Avoidance (HA), Reward Dependence (RD), and Persistence (P)— and three dimensions of character—Self-Directedness, Cooperativeness, and Self-Transcendence. In the present study, only the four temperament dimensions were considered.

#### Symptoms Check-list-90 (SCL-90) [24]

The SCL-90 is a 90-item self-report measure designed to assess a broad range of psychopathological symptoms. Higher scores indicate greater symptoms severity. In the present study, we considered only the Global Severity Index of the SCL-90 as a measure of overall psychopathological distress.

### 2.3 Data processing

The analysis begins from dividing the features derived from clinical data and pathological assessments, into a set of clustering features and a set of validating features. The clustering features constitute the space in which lie the data fed to the clustering algorithm of choice. The validating features constitute the space in which the separation of data performed by clustering can be statistically tested. This distinction is important, since every clustering algorithm explicitly or implicitly optimizes a measure of separation of dissimilar data or aggregation of similar data. Evaluating clustering results within the same feature space used for clustering can lead to overly optimistic conclusions. This is because clustering algorithms are inherently capable of partitioning data into groups, even when there is no true underlying structure or distinct generative processes. To assess the validity and relevance of the identified clusters, one can instead test the null hypothesis that cluster assignments are random with respect to a separate set of validating features i.e., features not used in the clustering process. If the cluster groupings exhibit statistically significant differences in these external validating features, this supports the validity of the clustering solution. Table 1 reports the 25 available features, categorized into clustering and validation features. For each feature, four summary statistics (minimum, mean, standard deviation and maximum) are provided, stratified by the three DSM-5 diagnostic groups. From the 25 features available, BMI, minimum BMI, weight suppression and amenorrhea, have been intentionally omitted from the clustering input. These variables are highly discriminative across DSM-5 diagnostic categories and would have led to a trivial separation of the bulimia nervosa cluster from anorexia nervosa subtypes. To ensure that the clustering process captured latent phenotypic structure beyond overt diagnostic distinctions, we chose to omit these features from the unsupervised model.

**Table 1:** Dataset Summary. Summary statistics for all dataset features, grouped according to the three DSM-5 diagnostic categories. Features are divided into those used for cluster definition and those reserved for validation. AN-R refers to anorexia nervosa, restricting type; AN-BP refers to anorexia nervosa binge/purge type; BN denotes bulimia nervosa; MD indicates mean; SD indicates standard deviation.

|  | Features | AN-R (351) |  |  | AN BP (150) |  |  | BN (308) |  |  |
| --- | --- | --- | --- | --- | --- | --- | --- | --- | --- | --- |
| | | min | MD ± SD | max | min | MD ± SD | max | min | MD ± SD | max |
| Clustering Features | Drive for thinness (DR-THIN) | 0 | 10.0 ± 6.8 | 21 | 0 | 14.4 ± 5.8 | 21 | 0 | 15.7 ± 5.1 | 21 |
| | Bulimia (BUL) | 0 | 2.0 ± 2.4 | 15 | 0 | 8.9 ± 5.7 | 21 | 0 | 12.9 ± 4.6 | 21 |
| | Body dissatisfaction (BODY-DISS) | 0 | 10.2 ± 6.8 | 27 | 0 | 13.3 ± 8.0 | 27 | 0 | 18.5 ± 7.6 | 27 |
| | Ineffectiveness (INEFF) | 0 | 8.2 ± 6.6 | 30 | 0 | 12.3 ± 7.8 | 30 | 0 | 10.9 ± 7.1 | 29 |
| | Perfectionism (PERF) | 0 | 4.7 ± 3.7 | 18 | 0 | 6.0 ± 4.1 | 17 | 0 | 6.1 ± 4.3 | 18 |
| | Interpersonal distrust (INT-DIS) | 0 | 6.6 ± 4.4 | 20 | 0 | 7.4 ± 4.8 | 21 | 0 | 6.7 ± 4.7 | 20 |
| | Interceptive awarness (INT-AW) | 0 | 8.2 ± 6.3 | 26 | 0 | 12.4 ± 6.7 | 29 | 0 | 12.2 ± 6.4 | 29 |
| | Maturity fears (MAT-FEAR) | 0 | 7.8 ± 4.6 | 24 | 0 | 8.8 ± 5.5 | 24 | 0 | 6.9 ± 4.6 | 29 |
| | Novelty seeking (NS) | 3 | 13.7 ± 4.8 | 29 | 5 | 16.8 ± 5.6 | 31 | 3 | 18.0 ± 5.6 | 31 |
| | Harm avoidance (HA) | 5 | 21.0 ± 6.3 | 21 | 3 | 21.5 ± 6.3 | 32 | 4 | 21.1 ± 6.4 | 34 |
| | Reward dependence (RD) | 3 | 12.3 ± 3.7 | 21 | 2 | 12.6 ± 3.9 | 20 | 1 | 13.1 ± 3.8 | 21 |
| | Persistence (P) | 1 | 6.3 ± 1.9 | 9 | 1 | 5.9 ± 2.1 | 9 | 0 | 5.4 ± 2.0 | 9 |
| | SCL90tot | 0 | 1.2 ± 0.7 | 3.1 | 0.2 | 1.6 ± 0.8 | 3.7 | 0.1 | 1.5 ± 0.7 | 3.6 |
| | tot ES | 0 | 8.9 ± 9.1 | 55 | 0 | 10.9 ± 8.4 | 41 | 0 | 11.4 ± 9.3 | 64 |
| | Age [years] | 12.7 | 20.8 ± 6.1 | 56.6 | 14.5 | 25.4 ± 7.6 | 57.0 | 15.2 | 24.4 ± 6.4 | 50.8 |
| | Age of Onset [years] | 12 | 17.8 ± 4.3 | 43 | 11 | 18.8 ± 6.0 | 53 | 8 | 17.9 ± 4.4 | 43 |
| | Illness Duration [years] | 0.3 | 2.0 ± 3.5 | 40 | 0.3 | 4.5 ± 4.8 | 26 | 0.3 | 4.4 ± 4.9 | 27 |
| | Physical Activity [hours/week] | 0 | 4.2 ± 4.3 | 48 | 0 | 4.0 ± 4.2 | 35 | 0 | 4.2 ± 4.2 | 30 |
|  | Pre-Adulthood Abuse (Yes) | 6.55% |  |  | 16.67% |  |  | 14.94% |  |  |
| Validation Features | Amenorrhea (Yes) | 90.31% |  |  | 67.33% |  |  | 11.69% |  |  |
| | BMI [kg/m<sup>2</sup>] | 9.4 | 15.9 ± 1.6 | 18.6 | 11.6 | 16.8 ± 1.3 | 18.4 | 18.11 | 22.8 ± 4.7 | 45.9 |
| | Minimum BMI [kg/m<sup>2</sup>] | 8.8 | 15.3 ± 1.7 | 18.6 | 11.2 | 15.5 ± 1.7 | 18.4 | 11.7 | 18.6 ± 3.2 | 44.1 |
| | Weight suppression BMI [kg/m<sup>2</sup>] | 0 | 23.6 ± 9.4 | 51.9 | 0 | 21.3 ± 10.4 | 54.5 | 0 | 9.6 ± 8.3 | 46.1 |
|  | Remission (No/Partial/Total) | 32.2%/34.5%/33.3% |  |  | 44.7%/30%/25.3% |  |  | 29.9%/21.8%/48.4% |  |  |
|  | DMS-5 Diagnosis (AN-R/AN-BP/BN) |  |  |  |  |  |  |  |  |  |

### 2.4 Pre-clustering analyses: diagnosis classification

Before applying the clustering procedure, which is an unsupervised approach, we first employed a supervised method to assess how well the DSM-5 diagnostic categories could be predicted from the clustering features described in subsection 2.3 and listed in Table 1.

XGBoost (eXtreme Gradient Boosting) is a widely used, high-performance ensemble learning algorithm, based on gradient-boosted decision trees. It iteratively constructs decision trees that correct errors made by previous ones, using gradient descent to minimize a specified loss function, with built-in regularization to mitigate overfitting. The dataset was split into 80% training and 20% testing subsets. To tune hyper-parameters, a grid search with 5-fold stratified cross-validation on the training set was used; the set of parameters tested is reported in the supplementary material section 1. A fixed seed (42) was randomly chosen to ensure reproducibility of the experiment. Due to class imbalance, the average balanced accuracy was used as the evaluation metric. By definition, for a multi-class classification problem with *C* classes (here the three DSM-5 diagnosis), the balanced accuracy is:

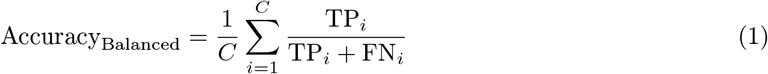

where TP_*i*_ and FN_*i*_ stands respectively for the true positive and the false negative for class *i*.

### 2.5 Clustering

Clustering is a popular unsupervised technique, particularly useful in the identification of phenotypes, which can be effectively used to find groups of patients with common traits. The following procedure was applied to cluster the data according to the selected features described in subsection 2.3.

1. The distance between subjects is calculated using the General Distance Measure (GDM) [25] on the 19 selected features. The GDM is a proximity measure inspired by Kendall’s General Correlation Coefficient, and thus apt both for numerical and ordinal scales. Between two subjects *i* and *j*, the distance *d*_*ij*_ is calculated.
2. The matrix *D*_(*ij*)_ = *d*_*ij*_ is obtained from the pairwise distances between patients. After that, the similarity matrix (*W* = 𝕀 − *D*) is calculated and used as input for the clustering algorithm.
3. The application of the Repeated Spectral Clustering (RSC) [26] algorithm to the similarity matrix *W* allows the identification of groups of patients with similar traits and common characteristics.

The RSC algorithm is a blend of spectral clustering [27], known to work well with similarities matrices representing graphs, and consensus clustering [28], known to be powerful since it allows to deal with the variability of results that stem from random initializations. In brief, RSC takes the matrix *W* as input to the spectral clustering procedure. Instead of running *k*-means [29] just once, it is performed *N*_*RSC*_ times to account for randomness in centroid initialization. Across these *N*_*RSC*_ iterations, the number of times two patients are assigned to the same cluster is recorded. The more frequently two subjects are clustered together, the stronger the evidence that they should remain in the same cluster in the final assignment. The final cluster assignment is based on the co-occurrence matrix *C*, where each element *C*_*ij*_ represents the number of times subjects *i* and *j* were clustered together across all runs. For a complete description of the algorithm, including technical details and its application to NIHSS scores, refer to Tshimanga and colleagues [26].

The optimal number of clusters is determined by calculating the maximum spectral gap for different cluster configurations (i.e. different *k*) [26]. The elbow criterion is a simple and effective way to find the optimal number of clusters. In particular, for *k* = 4, the maximum spectral gap is at the end of a high plateau, indicating well-separated groups, while for *k* = 5, it decreases sharply, indicating subjects that are worst separated in groups. Thus, *k* = 4 is chosen as the optimal number of clusters. For more details, see the supplementary material section 2.

### 2.6 Statistical tests

After applying the RSC clustering algorithm and determining the optimal number of clusters, statistical tests were conducted on the validation features to assess differences between the identified groups. The null hypothesis is that of no association between cluster label and diagnosis or remission, respectively. Both these categorical variables, have been compared between the four groups with the *χ*^2^ independence test [30] as omnibus and post hoc for pairwise comparisons, in order to compare the observed counts against the expectations under the null hypothesis. Expected frequencies are based on marginal distributions, assuming independence. Significant deviations between observed and expected distributions indicate non-random associations and support the clinical relevance of the identified clusters. The Cramér’s *V* metric [31] was used as effect size for the *χ*^2^ test; the significance level *α* was set to 0.05. While boolean and categorical features have been tested with the *χ*^2^ test, numerical and ordinal features (e.g BMI) between the four groups have been compared with the Kruskal-Wallis test [32] as omnibus, and then the Mann-Whiteny U [33] as post hoc for pairwise comparisons. Finally, p-values have been corrected for multiple comparisons using the Holm [34] procedure.

### 2.7 Post-clustering analyses: clusters classification

The XGBoost algorithm, using the same technical setup described in subsection 2.4, was applied to the clustering features selected in subsection 2.3 and listed in Table 1, this time to predict the cluster labels. Comparing the performance of predicting DSM-5 diagnoses versus predicting the cluster labels highlights the separability achieved by the RSC algorithm. This comparison also illustrates the extent to which the newly identified clusters, or phenotypes, are more strongly described by the selected clustering features.

## 3 Results

The RSC clustering procedure is applied to the 19 clustering features, giving an optimal number of clusters equal to 4. The maximum gap criterium, usually used in spectral-like clustering algorithms [26], suggests *k* = 4 as the optimal number of classes (see the supplementary material section 2).

### 3.1 Clusters and DSM-5 diagnosis

The partition of subjects into four clusters obtained via RSC is illustrated in Figure 1, Panel A. The visualization is based on the Fruchterman-Reingold force-directed layout algorithm [35], where the nodes connected by edges of higher weight are positioned closer together. The pairwise values of the edge weights correspond to the entries in the co-occurrence matrix *C*_*ij*_, which is the number of times two patients *i* and *j* have been clustered together over the *N*_*RSC*_ iterations performed by the RSC algorithm (see subsection 2.5). This representation highlights a high degree of cluster separability, with only six subjects occupying intermediate positions between clusters (IDs: 155, 271, 398, 692, 751 and 778). The same cluster layout is used in Figure 1 Panel B to illustrate the association between diagnostic categories and the identified clusters.

**Figure 1.**
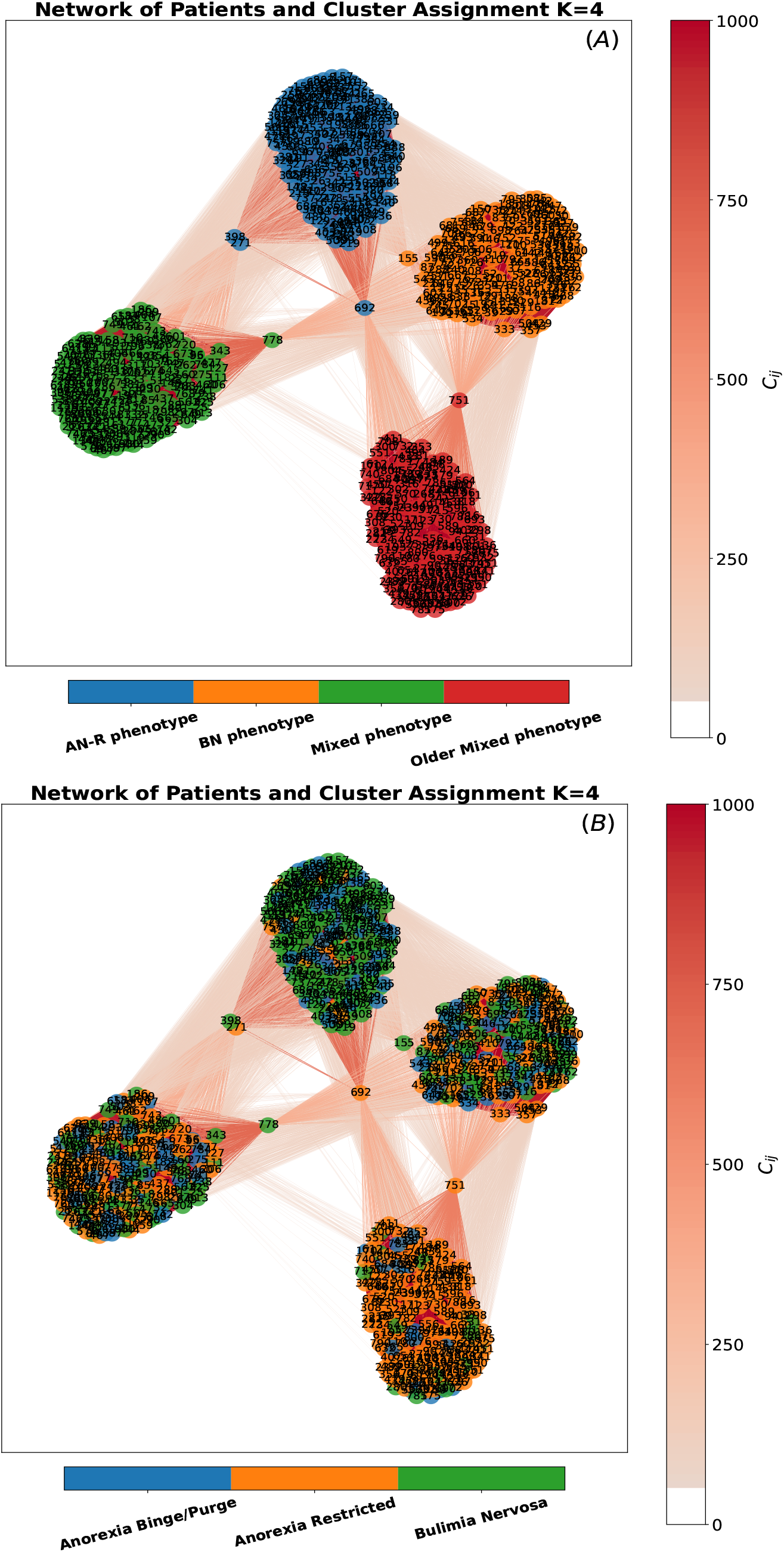
RSC results. Graph visualization of the clustering co-occurrence matrix *C*. Nodes represent patients; node numbers indicate patient IDs. In Panel A node colors identifies clusters, while in Panel B identifies one of the three DSM-5 diagnosis. Darker red and thicker links patients frequently clustered together. Weak links occurring in *<* 5% of repetitions are omitted.

The relationship between cluster assignments and both DSM-5 diagnoses and treatment remission was formally assessed using statistical tests of independence. This was done by comparing the observed frequencies of diagnostic and remission labels within each cluster to the expected frequencies under the assumption of random cluster assignment.

For diagnosis, the omnibus *χ*^2^ independent test turns out to be significant (*χ*^2^(6, 809) = 248, *p <*.001, *V* =.39), with strong effect size. The strongest associations were observed between Cluster 1 and restricting anorexia, and between Cluster 2 and bulimia nervosa. Specifically, 81.8% of individuals in Cluster 1 were diagnosed with restricting anorexia, while 67.6% of those in Cluster 2 were diagnosed with bulimia nervosa. Pairwise post hoc comparisons between these two clusters and all the others were highly significant. Finally, no significant differences were found between Cluster 3 and Cluster 4, both of which included a heterogeneous mix of subjects with different diagnoses.

Concerning remission, the omnibus *χ*^2^ independent test turns out to be significant (*χ*^2^(6, 809) = 13.9, *p* =.03, *V* =.09), but with weak effect size. Although none of the pairwise post hoc comparisons reached statistical significance, a descriptive pattern emerged: Cluster 1 was characterized by a higher proportion of partial remission (36.5%) and a lower rate of full remission, whereas Cluster 2 showed the opposite trend, with more individuals achieving full remission (44.7%) and fewer cases of no or partial remission. Clusters 3 and 4 displayed a more heterogeneous pattern, with few cases of partial remission and a comparable number of patients in full or no remission.

### 3.2 Phenotypes of the clusters

Each cluster has a characteristic profile across the input clustering features. By design, the clustering algorithm yields a partition that separates the data across features. In Table 2 it is reported a summary table describing the four clusters, respect the 25 features, divided in clustering and validation features.

Cluster 1 (AN-R phenotype) is composed predominantly (81.8%) by patients with restricting-type anorexia nervosa. Individuals in this group are younger than those in other clusters (18.2 ± 3.6 years), with an adolescent age of onset (15.7 ± 2.1 years), short illness duration (1.4 ± 2.3 years), markedly low BMI (16.4 ± 2.2) and minimum BMI (15.7 ± 1.8). This cluster also show the lowest levels of self-reported psychopathology, both general and ED-specific, across the sample. From a temperamental standpoint, patients in this cluster exhibit the lowest level of harm avoidance (17.7 ± 5.7). Individuals in this cluster also report the lowest number of stressful life events (6.8 ± 7.0) and experiences of childhood abuse (3.12%).

Cluster 2 (BN phenotype) is composed predominantly (70.7%) of patients diagnosed with bulimia nervosa. Consistently with their diagnostic profile, individuals in this group show the highest scores on the EDI bulimia scale (12.2 ± 4.7), as well as higher current BMI (21.2 ± 4.9) and minimum lifetime BMI (17.8 ± 3.3). Notably, the latter two variables had not been explicitly included in the clustering

**Table 2.**
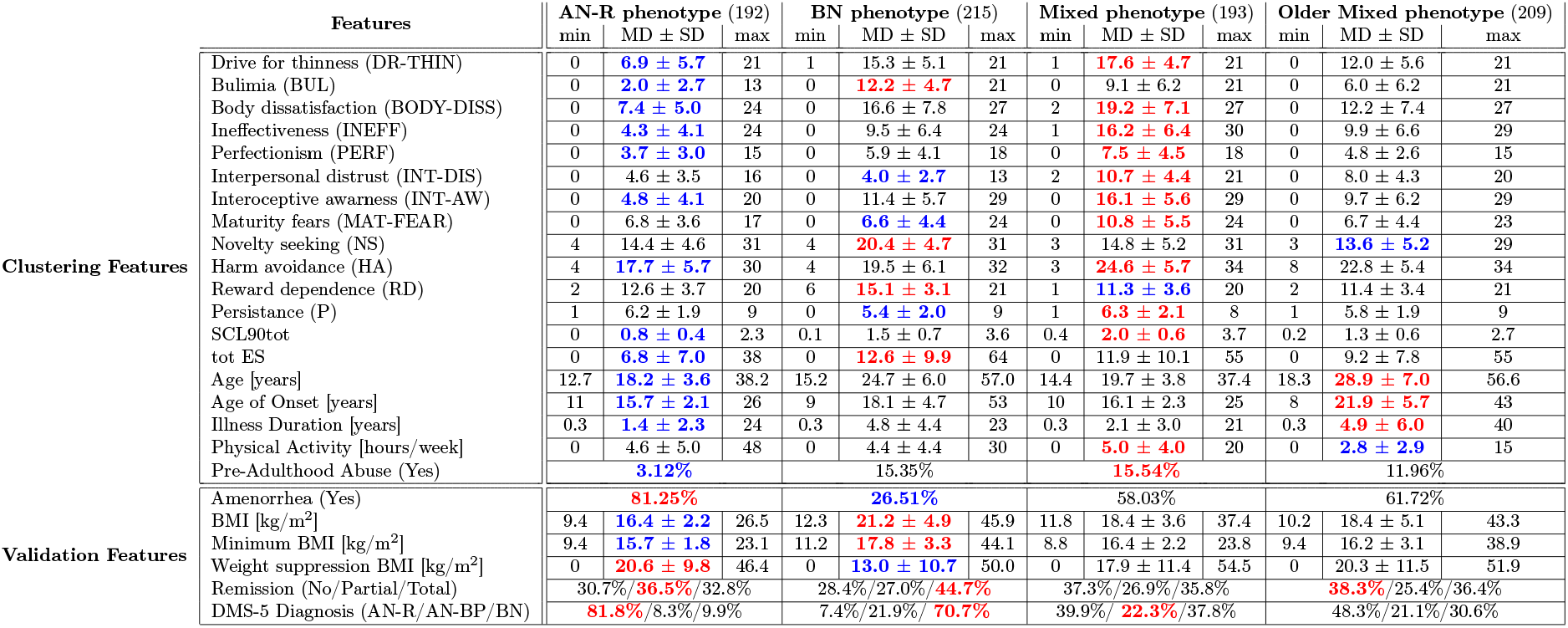
RSC Clusters Summary. Summary statistics for all dataset features, grouped according to the four clusters found by the RSC algorithm. Features are divided into those used for cluster definition and those reserved for validation. AN-R refers to anorexia nervosa, restricting type; AN-BP refers to anorexia nervosa binge/purge type; BN denotes bulimia nervosa; MD indicates mean; SD indicates standard deviation. The cluster with the highest mean value for each feature is highlighted in red, while the lowest is shown in blue. analysis, further supporting the meaningfulness of the identified groups. Regarding temperamental traits, the individuals in this group display the highest levels of novelty seeking (20.4 ± 4.7) and reward dependence (15.1 ± 3.1) and the lowest level of persistence (5.4 ± 2.0). This profile suggests a tendency toward impulsivity and a low tolerance for frustration. This cluster also show the highest rates of stressful life events (12.6 ± 9.9).

Cluster 3 (Mixed phenotype) include a diagnostically mixed sample of patients-comprising individuals with restrictive anorexia nervosa (39.9%), binge-purging anorexia nervosa (22.3%), and bulimia nervosa (37.8%). It is characterized by the highest levels of psychopathology, encompassing both eating disorder–specific and general psychiatric symptoms. Specifically, it shows the highest average scores across the entire EDI inventory (excluding the Bulimia subscale) and the SCL-90 (2.0 ± 0.6), compared to all other clusters. Regarding temperamental traits, the individuals in this cluster present the lowest level of reward dependence (11.3 ± 3.6) and the highest levels of harm avoidance (24.6 ± 5.7) and persistance (6.3 ± 2.1). This group also shows the highest prevalence of childhood trauma (15.54%) and recent stressful life events (11.9 ± 10.1). Patients in this group also exhibit the highest average physical activity (5.0 ± 4.0 hours/week).

Cluster 4 (Older Mixed phenotype) includes patients with a mixed phenotype, as both anorexia nervosa (69.4%) and bulimia nervosa (30.6%) diagnoses are represented. Compared to the other clusters, this group is characterized by older age (28.9 ± 7.0 years), later illness onset (21.9 ± 5.7 years), and longer illness duration (4.9 ± 6.0 years). Psychopathology levels are intermediate relative to the other subgroups and the temperamental profile is marked by the lowest level of novelty seeking (13.6 ± 5.2 years). This group also shows the highest proportion of patients who did not achieve remission (38.3%).

The distribution of values for each feature and cluster is shown in Figure 2. To ensure comparability across all features, each variable was rescaled using min-max normalization to the range [0, 1]. For features with well-defined bounds (e.g. those from EDI and TCI), normalization was performed using their known minimum and maximum values. In contrast, features without predefined bounds (e.g. age, BMI etc.) were normalized using the empirical minimum and maximum observed inside the cohort. For boolean variables (e.g. amenorrhea) and categorical variables (e.g. remission), a stacked bar plot was used to visualize their distribution across clusters. The height of each segment within a bar represents the percentage of occurrences for each category within the corresponding cluster.

**Figure 2.**
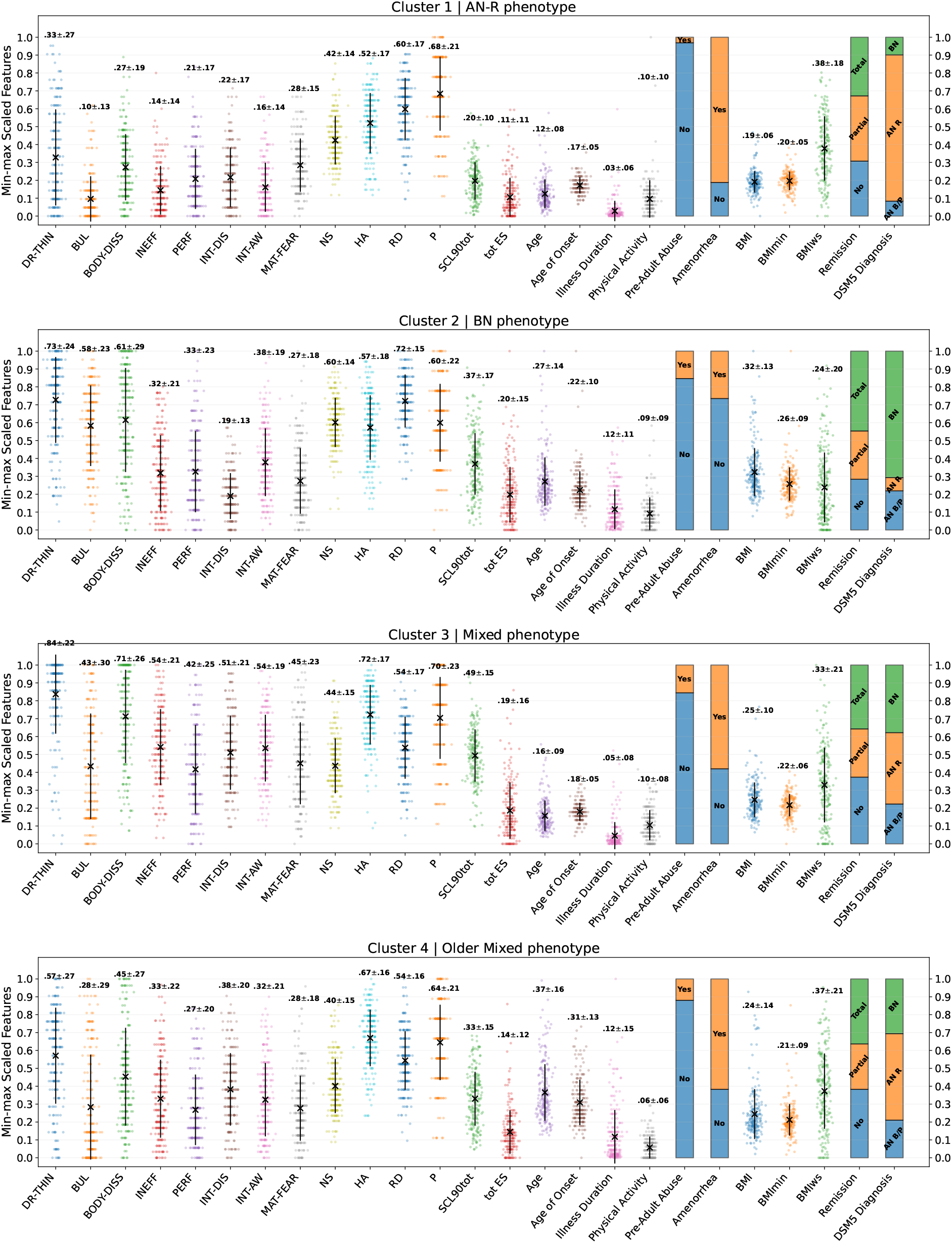
Clustering phenotypes. Each panel corresponds to one cluster identified by the RSC algorithm. The x-axis lists the available features—those used for clustering appear on the left, while validation features are shown on the right. All features are normalized to the 0–1 range for comparability. The plotted values represent the mean (cross) and standard deviation (line) for each feature. Individual data points represent subjects.

### 3.3 Supervised learning comparison

A supervised learning analysis using XGBoost [36] was conducted prior to clustering to evaluate whether the 19 selected features could accurately predict DSM-5 diagnostic categories. The model, appropriately trained and tuned, achieved a balanced accuracy of 66.1%, indicating only moderate classification performance. This limited predictive power suggests that the selected features do not fully capture the heterogeneity inherent in the DSM-5 diagnoses.

In contrast, when predicting the data-driven cluster labels derived from the RSC algorithm, the same XGBoost model, appropriately trained and tuned, achieved a balanced accuracy of 92.4%, indicating great classification performance. This substantial improvement highlights that while both unsupervised and supervised approaches struggle to replicate DSM-5 groupings, they may converge on similar discriminative patterns that support more robust classification when alternative, data-driven phenotypes are employed. The optimal hyper-parameters found for both tasks are reported in the supplementary material section 1.

## 4 Discussion

In this study, an unsupervised machine learning approach was applied to a large and phenotypically heterogeneous sample of individuals with EDs, yielding four distinct clusters. Two of these clusters demonstrated a high degree of alignment with established DSM-5 diagnostic categories, specifically, patients with anorexia nervosa restricting-type and those with bulimia nervosa. In contrast, the remaining two clusters reflected more complex and diagnostically mixed phenotypes, suggesting the presence of latent clinical subtypes not fully captured by current nosological systems. This pattern of findings is consistent with the overarching aim of the study: to examine the extent to which data-driven classifications converge with, or diverge from, traditional categorical diagnoses. On one hand, the emergence of diagnostically coherent clusters reaffirms the clinical validity of existing DSM-5 constructs. On the other, the identification of diagnostically heterogeneous subgroups highlights the phenotypic complexity and dimensional variability that characterize eating disorders, an aspect which is increasingly recognized in contemporary psychiatric research [37]. A noteworthy feature of this study is the nature of the input data. The clustering procedure relied exclusively on self-report instruments that can be easily employed in clinical practice, complemented by easily accessible clinical and demographic variables. This highlights the pragmatic potential of clustering approaches, which can be implemented using data commonly available in real-world settings [38]. Such findings support the notion that data-driven, ML-based phenotyping may offer a cost-effective and scalable strategy for enhancing diagnostic precision, guiding personalized treatment, and informing risk stratification. Future research should aim to refine these models and evaluate their integration into clinical decision-making pathways, with particular attention to their ability to delineate clinically meaningful profiles that extend beyond conventional diagnostic boundaries.

### 4.1 Phenotypes and targeted intervention

Beyond validating the effectiveness of the clustering approach, the present findings revealed four phenotypically distinct subgroups, each characterized by a specific configuration of demographic, psychopathological, and temperamental features. These profiles allow for clinically grounded considerations regarding the phenotypic variability of EDs and their potential implications for diagnostic refinement and therapeutic planning. Among the four clusters, Cluster 1 and Cluster 2 demonstrated the highest degree of alignment with categorical DSM-5 diagnoses; Clusters 3 and 4, in contrast, showed greater diagnostic heterogeneity and phenotypic complexity, and did not correspond as clearly to established diagnostic categories.

Cluster 1 (AN-R phenotype) is composed predominantly by patients with restricting-type anorexia nervosa. The clinical characteristics described in subsection 3.2 and in Table 2 are indicative of an early-stage presentation of the disorder, marked by significant physical severity (e.g. low BMI and amenorrhea). Despite this physical severity, individuals in this cluster report relatively low levels of psychopathology. These features, combined with young age and short illness duration, are consistent with previous findings indicating that, in the early stages of the disorder, egosyntonic symptoms and limited insight may attenuate subjective distress [39]. Such a profile may have therapeutic relevance, supporting the use of interventions aimed at enhancing illness awareness, strengthening motivation for change, and improving engagement in treatment tailored to restrictive anorexia nervosa.

Cluster 2 (BN phenotype) is composed predominantly of patients diagnosed with bulimia nervosa. The clinical and psychological profile of this cluster reflects a pattern consistent with an affective–impulsive subtype previously described in eating disorder populations, which is characterized by the presence of emotional instability, impulsivity, and lower interpersonal avoidance. The notably higher rate of full remission observed in this cluster suggests a relatively favorable treatment response. These findings are in line with prior literature indicating that individuals with similar profiles may particularly benefit from structured, symptom-focused interventions [40], potentially accounting for the more positive outcomes seen in this group.

Cluster 3 (Mixed phenotype) was defined by the highest burden of psychopathology, encompassing both ED-specific and general psychiatric symptoms. The combination of diagnostic heterogeneity and severe psychopathology, suggests a transdiagnostic profile marked by broad dysregulation across emotional, identity-related, and interpersonal domains. The elevated rates of trauma and life stressors are consistent with prior evidence linking trauma exposure to greater clinical complexity in EDs [41]. From a temperamental point of view, this cluster is characterized by avoidant and rigid personality traits, frequently described in patients with anorexia nervosa. These features are well documented in the literature and may also have relevant therapeutic implications [42]. Clinically, this subgroup may benefit from integrated, multimodal interventions that target both ED-specific symptoms and broader comorbid dimensions, such as affective instability, cognitive rigidity, and trauma-related distress.

Cluster 4 (Older Mixed phenotype) includes patients with different diagnoses, characterized by a combination of older age, longer illness duration, and moderate psychopathology. These aspects suggest a more chronic and potentially stabilized clinical presentation. The presence of both anorexia nervosa and bulimia nervosa diagnoses, along with a controlling–compulsive temperament, points to a heterogeneous but less acutely severe profile. These characteristics align with patterns of persistent impairment and may indicate the need for long-term, rehabilitative treatment approaches focused on functional recovery and symptom management rather than short-term resolution.

One of the aims of this study was to explore whether the data-driven phenotypic clusters were meaningfully associated with clinical outcome, defined categorically as full remission, partial remission, or no remission. Although the overall distribution of outcomes appeared relatively balanced across clusters, statistical testing rejected the hypothesis of independence, suggesting that remission status is not randomly distributed with respect to cluster assignment. Notably, Cluster 2 showed the highest proportion of full remission cases. In contrast, Cluster 1, largely composed of individuals with restricting-type anorexia nervosa, was more frequently associated with partial remission. Clusters 3 and 4, both marked by diagnostic heterogeneity and prolonged illness duration, displayed a more unfavorable outcome profile. In particular, Cluster 4 had the highest proportion of individuals who did not reach remission, pointing to a potentially more chronic and treatment-resistant trajectory. While these patterns did not reach significance in pairwise comparisons, they may reflect clinically relevant trends that warrant further investigation. The categorical nature of the remission variable and the cross-sectional assessment limit definitive conclusions. Future studies with longitudinal designs and more nuanced outcome indicators, such as relapse rates, psychosocial functioning, or quality of life, will be necessary to assess the prognostic value of these phenotypic distinctions more robustly.

### 4.2 Toward a transdiagnostic understanding of EDs

Beyond the formal clustering structure, our findings contribute meaningfully to the growing body of literature advocating for a dimensional and transdiagnostic reconceptualization of EDs. While two of the identified clusters closely aligned with prototypical diagnostic categories, restricting-type anorexia nervosa and bulimia nervosa, the remaining two clusters revealed more heterogeneous profiles, marked by overlapping symptomatology and divergent profiles. This duality illustrates a key point: although established diagnostic constructs retain clinical validity for a subset of patients, they may obscure important variability in others whose presentation is not adequately captured by current nosological boundaries. The identification of diagnostically mixed clusters speaks to the multifaceted nature of transdiagnostic complexity. Importantly, this complexity may take at least two distinct forms.

The first may be considered a form of cross-sectional transdiagnosticity, exemplified by individuals who simultaneously exhibit elevated levels of general psychopathology, emotional dysregulation, and trauma-related vulnerability. These cases may reflect a shared vulnerability profile cutting across EDs subtypes, consistent with models that emphasize impairments in affect regulation, identity coherence, and interpersonal functioning as common etiological threads [43, 44].

The second may represent a form of temporally diffuse transdiagnosticity, as suggested by the presence of individuals with older age and prolonged illness duration. It is possible to speculate that this pattern aligns with the phenomenon of diagnostic migration described in longitudinal studies, and may reflect the cumulative transformation or substitution of symptoms over time within the spectrum of ED [45]. These two dimensions of transdiagnosticity highlight the limitations of static diagnostic systems in fully capturing the evolving nature of eating disorders.

Although categorical diagnoses can be informative at specific time points, they often do not account for symptom progression, accumulation of comorbidities, and resistance to treatment. In this regard, data-driven approaches, such as the one employed in this study, may help uncover latent phenotypes that better reflect both the underlying mechanisms and the real-world complexity of these conditions. Moreover, from a clinical standpoint, these findings suggest that different phenotypic profiles may call for distinct therapeutic strategies. Individuals belonging to diagnostically coherent clusters may respond well to targeted, diagnosis-specific interventions. In contrast, patients with mixed or complex profiles may require broader, integrative approaches that target transdiagnostic mechanisms such as emotion regulation or behavioral rigidity, and that can be flexibly adapted over time. Moreover, the ability to empirically distinguish between stable, early-stage phenotypes and long-standing, treatment-resistant trajectories offers a promising avenue for improving treatment stratification and personalizing care pathways. Ultimately, this study reinforces the relevance of transdiagnostic models in both research and clinical practice. By identifying subgroups that are not fully captured by current diagnostic labels, it supports the need for classification systems that can integrate both categorical and dimensional information, reflect clinical reality, and guide the development of mechanism-based interventions tailored to individual needs.

### 4.3 Strengths and limitations

This study presents some important strengths. First, it is based on a large, phenotypically diverse sample of patients with eating disorders, all assessed at a single specialized center using standardized diagnostic and psychometric procedures. This uniformity in data acquisition minimizes methodological heterogeneity and enhances the internal validity of the findings. Second, the application of a novel unsupervised machine learning pipeline represents a methodological innovation within the field. Unlike more commonly used clustering techniques, this approach enhances robustness and interpretability, allowing for the identification of coherent phenotypic subgroups without relying on a priori diagnostic assumptions. The fact that these clusters emerged from easily accessible self-report and clinical data further underscores the potential for translational applications in both research and practice.

Despite its strengths, some limitations of the research must also be acknowledged. First, the crosssectional design precludes any inference about the temporal stability or prognostic value of the identified phenotypes. Longitudinal data would be necessary to determine whether these clusters remain stable over time or evolve in response to treatment and illness progression. Second, the study sample consisted exclusively of female patients, limiting the generalizability of findings to male or gender-diverse populations, who may exhibit different symptom profiles, risk factors, and trajectories of illness. Third, while the use of standardized self-report instruments enhances the scalability of the clustering procedure, it also introduces potential biases related to self-perception and reporting accuracy.

Behavioral or biological markers were not included and could enrich the phenotypic profiles in future studies. Finally, although the ML pipeline yielded clinically interpretable clusters, its application to other independent samples is important to evaluate reproducibility, generalizability, and clinical utility. At present, the integration of such unsupervised models into real-world clinical workflows remains exploratory, and further research is needed to determine whether these tools can meaningfully inform diagnostic decision-making, treatment planning, or prognostic stratification in diverse clinical settings.

## 5 Conclusions

In recent years, machine learning have advanced significantly, offering new prospects across many domains, including eating disorders. Despite this promise, existing ML applications to these clinical populations have been somewhat fragmented. Notably, many of the existing studies remain anchored in traditional diagnostic labels, and fewer have fully explored unsupervised methods. In this paper, we employed RSC to a large cohort of female patients with eating disorders, relying on self-report and clinical variables. This approach yielded four robust and clinically interpretable clusters. Two of these aligned with DSM-5 prototypical diagnoses, supporting their empirical validity. However, the other two revealed diagnostically mixed profiles marked by greater psychopathological burden, prolonged illness duration, or trauma exposure. These findings underscore the value of unsupervised methods for uncovering latent phenotypic structures that may transcend conventional nosological boundaries. From a clinical perspective, our results suggest that patients fitting categorical profiles may benefit from targeted interventions, whereas those with complex or overlapping features may require more integrative and flexible treatment strategies. Overall, this work illustrates how data-driven phenotyping can inform a more nuanced, stratified understanding of EDs, ultimately contributing to the development of personalized and scalable models of care.

## Data Availability

All data produced in the present study are available upon reasonable request to the authors.

## 6 Acknowledgment

## Funding

This work was supported by the STARS@UNIPD funding program of the University of Padova, Italy, through the project: MEDMAX. This project has received funding from the European Union’s Horizon Europe research and innovation programme under grant agreement no 101137074 - HEREDITARY.

## Conflict of interest

The authors have no competing interests to declare that are relevant to the content of this article.

## Code availability

The code is available at MedMaxLab/clusteringED.

## Data availability

The data that support the findings of this study are available upon reasonable request.

## Ethical statement

The research was conducted in accordance with the principles embodied in the Declaration of Helsinki and in accordance with local statutory requirements. All participants (or their parent or legal guardian in the case of children under 18) gave written informed consent to participate in the study. All subjects gave their informed written consent to the use of data in an anonymous form. The Ethical Committee of Padova Hospital approved the study (protocol n*◦* 1598P).

## Authors’ contributions

AZ - Conceptualization, Methodology, Statistical Analysis, Writing - Original Draft. LFT - Methodology, Writing - Original Draft. VM - Medical Interpretation, Writing - Original Draft. AF - Conceptualization, Medical Interpretation, Supevision. MA - Conceptualization, Methodology, Project Administration, Supervision, Writing - Original Draft. EC - Conceptualization, Medical Interpretation, Supervision, Writing - Original Draft. All authors reviewed and edited previous versions of the manuscript.

## Notes

### Competing Interest Statement

The authors have declared no competing interest.

### Author Declarations

The Ethical Committee of Padova Hospital gave ethical approval for this work (protocol n. 1598P).

