## Supplementary Material for "Stratifying Eating Disorders with Clustering: From Diagnosis to Phenotypic Diversity"

### 1 Supplementary Material

#### 1.1 XGBoost Optimization

In this section, we report the hyperparameter grid search performed to optimize the XGBoost classifier. Two supervised tasks were conducted: one in which the model predicted the DSM-5 diagnosis, and another where the model predicted the clustering labels previously identified. The optimal hyperparameter values obtained are summarized in Table 1.

| **Hyper-Parameter** | **Search Space** | **Optimal per DSM5** | **Optimal per Label** |
|---|---|---|---|
| Number of estimators | [50, 100, 200] | 100 | 200 |
| Maximum tree depth | [3, 6, 12] | 6 | 12 |
| Learning rate | [0.01, 0.1, 1] | 0.1 | 0.1 |
| Minimum child weight | [0.5, 1, 2] | 0.5 | 2 |
| Sub-sample | [0.1, 0.5, 1] | 0.5 | 0.5 |
| Col-sample by tree | [0.1, 0.5, 1] | 0.5 | 0.5 |

Table 1: Grid of hyper-parameters used for XGBoost tuning, with the best values found for the two supervised task considered.

#### 1.2 Optimal Number of Clusters

The way in which $K$ is chosen, relies on the spectra of the normalized Laplacians of the matrices $W, C$. In classical spectral clustering, $n$ null eigenvalues indicate $n$ completely separate groups in the data graph, and $n$ very small eigenvalues followed by a larger eigenvalue indicate $n$ softly distinct groups. The max gap criterion would suggest to choose $k$ for a given graph so that the $k$-th spectral gap (the difference between $k+1$-th and $k$-th eigenvalues) is maximal. The actual choice must be made among the different max gaps across the matrices $C_k$, for every $k$ attempted. In Figure 1, it is reported the max-gap, for various $k$.

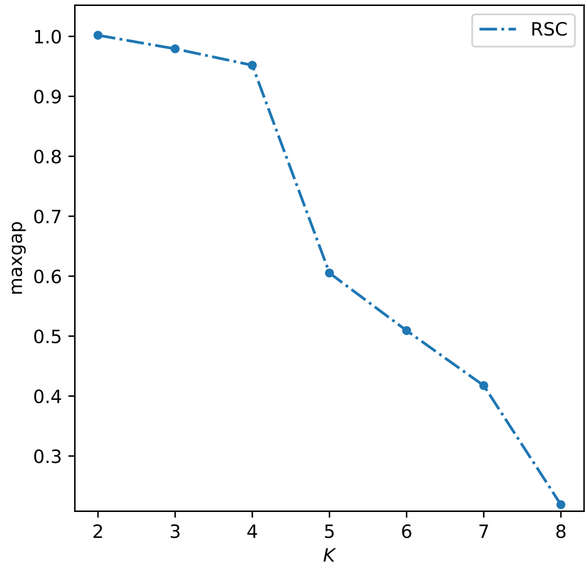


Figure 1: Max-gap statistic computed from the RSC algorithm for different values of $K$. A high plateau is observed up to $K = 4$, indicating well-separated clusters. From $K = 5$ onward, the statistic drops sharply, suggesting reduced cluster separability. This pattern indicates that $K = 4$ represents a good trade-off between model complexity and the quality of the clustering solution.
